# A randomised crossover trial of high- versus low-protein diets on glycaemia in people with normal glucose tolerance (NGT) and type 2 diabetes (T2D)

**DOI:** 10.1101/2025.06.24.25330146

**Authors:** Nicola D Guess, Victoria Adejare, Nick Oliver, Richard Stevens, Jimmy D Bell

## Abstract

**Objective:** To determine whether a low-carbohydrate, high-protein diet lowers glucose compared to a low-carbohydrate, normal-protein diet.

**Research Design and Methods:** A total of 11 participants with type 2 diabetes (T2D) and 9 with normal glucose tolerance (NGT) were prescribed diets high (30%kcal) and low (15%kcal) in protein with carbohydrate kept constant (20%kcal), in randomised order (high-protein > low-protein > high-protein or low-protein > high-protein > low-protein sequence) following an initial run-in period of 1-2 days where participants followed their habitual diet (protein 15kcal; carbohydrate ∼45%kcal). Glucose was measured using blinded continuous glucose monitoring

**Results:** In people with T2D, 24-hour and post-prandial AUC □ □ □ were significantly lower (- 1.0mmol/L, 95%CI: -1.2 to -0.8, and (381.4 mmol/L*180, 95%CI: -443.5 to -319.3) and time in range (TIR) significantly higher (13.9%, 95%CI: 10.0 to 17.8) for the high-protein condition, compared to the low-protein condition. For people with NGT only post-prandial sensor glucose was lower on the high-protein compared to low-protein condition.

**Conclusion:** A high-protein, reduced-carbohydrate diet could be useful as a standalone or an adjunct to weight loss to help achieve optimum glucose concentrations for people with T2D.

Trial registration and protocol: ISRCTN26058700.

## Background

The degree of weight loss required to achieve remission of type 2 diabetes (T2D) may limit its applicability to many people with T2D across the population, and strategies to lower blood glucose independent of weight would be valuable [1]. In well-controlled studies, low-carbohydrate diets (20-40% Kcal from carbohydrate) lower glucose without weight loss, but the protein in the low-carbohydrate group was also significantly increased (30%kcal protein) compared to the high-carbohydrate group (15%kcal protein)[2, 3]. Emerging evidence suggests that reducing carbohydrate without increasing protein does not lower glucose, but this has never been tested [4, 5].

The objective of this study was to determine whether a low-carbohydrate diet with added protein lowers glucose compared to a low-carbohydrate diet without added protein.

### Research Design and Methods

In this randomised crossover trial, 12 people with and 9 people without T2D, recruited via social and print media advertisement were randomised to receive diets high (30%kcal) and low (15%kcal) in protein with carbohydrate kept at 20%kcal, following an initial controlled run-in period of 1-2 days where participants followed their usual low-protein (15kcal), high-carbohydrate (∼45%kcal) diet.

The idea for the study arose from patient and public involvement (PPI) groups who wanted “weight neutral” ways to manage their diabetes. The PPI groups also helped design the meal plans.

Participants were recruited using social media, screening and support visits took place at the Physiology Unit at the University of Westminster in London, United Kingdom, or remotely.

The inclusion criteria for all participants were: males and post-menopausal females, aged 45 to 60 years with a BMI of 23 to 35kg/m^2^ reporting a stable weight for 3 months prior to study commencement and without kidney disease, type 1 diabetes, liver disease, hematologic abnormalities, congestive heart failure, or untreated thyroid disease. Participants with T2D had to be diagnosed within the last 3 years and be diet-controlled or on metformin. If fasting plasma glucose exceeded 10mmol/L on two consecutive days the participants would be withdrawn from the study.

Participants were given individual meal prescriptions comprising three main meals and one snack to maintain their body weight, using the NIH meal planner [6], and were asked to weigh themselves once every morning and to complete a daily checklist of protein, and carbohydrate foods eaten to promote compliance.

The original study was designed without washouts between the two- and one-week dietary assignments, however due to logistical challenges with our first participant (out of country travel) we introduced wash-out periods of at least two weeks between dietary assignment. Randomisation was done using the free tool Research Randomiser. The investigators nor the participants were blinded to dietary assignment.

Sensor glucose was measured using a blinded Dexcom G6 continuous glucose sensor during 1-2 days of the “usual diet”, and during each of the three dietary assignments.

The primary outcome was the 24-h sensor glucose concentrations (mean of all interstitial glucose values over the final 5 days of each dietary assignment). Sample size was determined based on an expected -0.5mmol/L reduction in the high-protein compared to the low-protein group with a SD of 0.45 at 90% power and an alpha of 0.05 which required 11 participants. We aimed to recruit 15 to account for dropouts. Secondary outcomes included fasting sensor glucose (mean of all interstitial glucose values 2-hour prior to breakfast) and post-prandial sensor glucose concentrations, (tAUC_glucose_ 180 minutes after meal commencement) and time in range (TIR)(% of time spent between 3.9 and 10mmol/L). Data were analysed using mixed models with participant as fixed, time as main effect and randomization as covariate. The study was approved by the research ethics committee of the University of Westminster.

### Results

Following telephone screening of n=126 people, n=17 people with T2D and n=16 with NGT met the initial inclusion criteria, and n=12 T2D and n=12 NGT were randomised. In total n=11 T2D and n=9 NGT participants completed the trial (See Supp data for CONSORT diagram). Table 1 summarises the baseline participant characteristics.

**Table 1:**
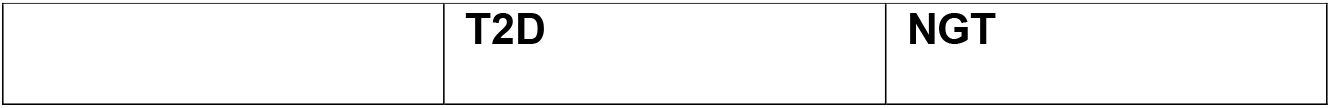
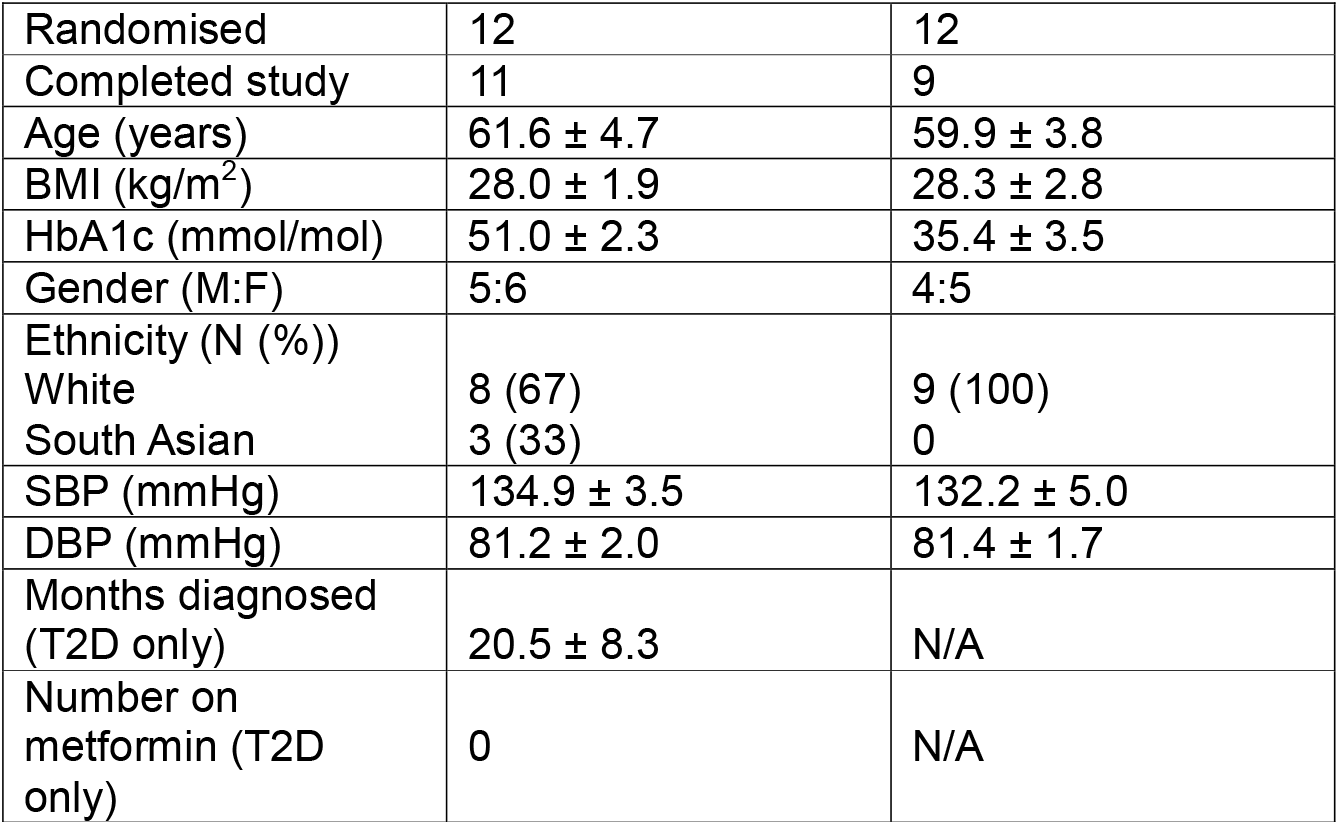
Baseline demographics and clinical data for T2D and NGT participants. NGT = normal glucose tolerance; T2D = type 2 diabetes. Values are mean ± SD.

|  |  |  |
| --- | --- | --- |
|  | **T2D** | **NGT** |
| Randomised | 12 | 12 |
| Completed study | 11 | 9 |
| Age (years) | 61.6 ± 4.7 | 59.9 ± 3.8 |
| BMI (kg/m<sup>2</sup>) | 28.0 ± 1.9 | 28.3 ± 2.8 |
| HbA1c (mmol/mol) | 51.0 ± 2.3 | 35.4 ± 3.5 |
| Gender (M:F) | 5:6 | 4:5 |
| Ethnicity (N (%)) |  |  |
| White | 8 (67) | 9 (100) |
| South Asian | 3 (33) | 0 |
| SBP (mmHg) | 134.9 ± 3.5 | 132.2 ± 5.0 |
| DBP (mmHg) | 81.2 ± 2.0 | 81.4 ± 1.7 |
| Months diagnosed (T2D only) | 20.5 ± 8.3 | N/A |
| Number on metformin (T2D only) | 0 | N/A |

In people with T2D (n=11), 24h glucose (Unadjusted mean change: -1.0mmol/L, 95%CI: -1.2 to -0.8), Figure 1) and AUC_180mins_ (Unadjusted mean change: (381.4 mmol/L*180mins, 95%CI: -443.5 to -319.3), Figure 2) were significantly lower during the high-protein condition compared to the low-protein condition, and TIR (Unadjusted mean change: 13.9%, 95%CI: 10.0 to 17.8, Figure 2) higher in the high-protein condition compared to the low-protein condition at all timepoints. After adjustment for baseline imbalance the difference in fasting glucose was not significantly different (Table 2). In people with NGT (n=9), the AUC*180mins was significantly lower in the high-protein condition compared to the high-protein condition at all timepoints, but there was no difference in 24h glucose or fasting glucose (Table 2).

**Table 2:** *The treatment effects after adjustment for baseline imbalance. Group A was randomized to high-protein first, and Group B was randomized to low-protein first. AUC (area under the curve); TIR = time in range.

|  | Timepoint | Estimate (Group A-Group B) | 95% Confidence Interval |  | Sig |
| --- | --- | --- | --- | --- | --- |
|  |  |  | Lower | Upper |  |
| **T2D (n=11)** |  |  |  |  |  |
| 24h sensor glucose (mmol/L) | 1 | -1.1 | -1.8 | -0.4 | 0.002 |
|  | 2 | 0.85 | 0.2 | 1.5 | 0.014 |
|  | 3 | -1.2 | -1.8 | -0.5 | 0.001 |
| TIR (%) | 1 | 15.5 | 5.1 | 25.8 | 0.005 |
|  | 2 | -12.1 | -22.5 | -1.8 | 0.0023 |
|  | 3 | 15.9 | 5.5 | 26.2 | 0.004 |
| AUC (mmol/L*180 mins) | 1 | -404.7 | -571.0 | -238.5 | <.001 |
|  | 2 | 324.6 | 158.4 | 490.8 | <.001 |
|  | 3 | -456.3 | -622.5 | -290.1 | <.001 |
| Fasting glucose (mmol/L) | 1 | -0.7 | -1.3 | -0.01 | 0.05 |
|  | 2 | 0.6 | -0.1 | 1.2 | 0.072 |
|  | 3 | -0.7 | -1.3 | -0.04 | 0.04 |
| **NGT (n=9)** |  |  |  |  |  |
| 24h sensor glucose | 1 | -0.5 | -1.0 | 0.1 | 0.084 |
|  | 2 | 0.5 | -0.0 | 1.0 | 0.069 |
|  | 3 | -0.4 | -1.0 | 0.2 | 0.156 |
| AUC (mmol/L*180 mins) | 1 | -181.6 | -316.8 | -46.3 | 0.01 |
|  | 2 | 180.4 | 45.1 | 315.7 | 0.011 |
|  | 3 | -174.4 | -309.6 | -39.1 | 0.013 |
| Fasting glucose (mmol/L) | 1 | -0.2 | -0.8 | 0.5 | 0.596 |
|  | 2 | 0.5 | -0.2 | 1.1 | 0.165 |
|  | 3 | -0.2 | -0.9 | 0.5 | 0.487 |

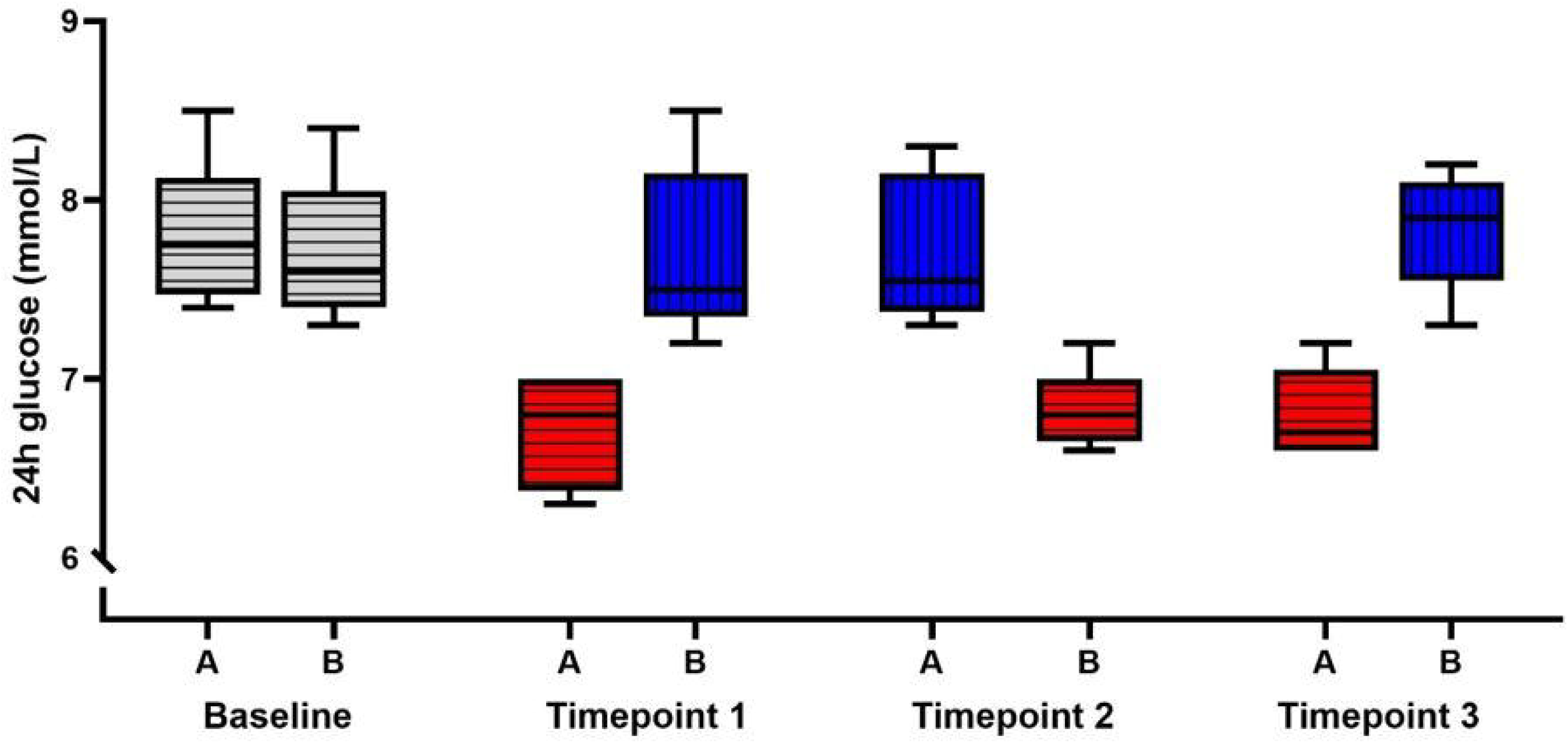

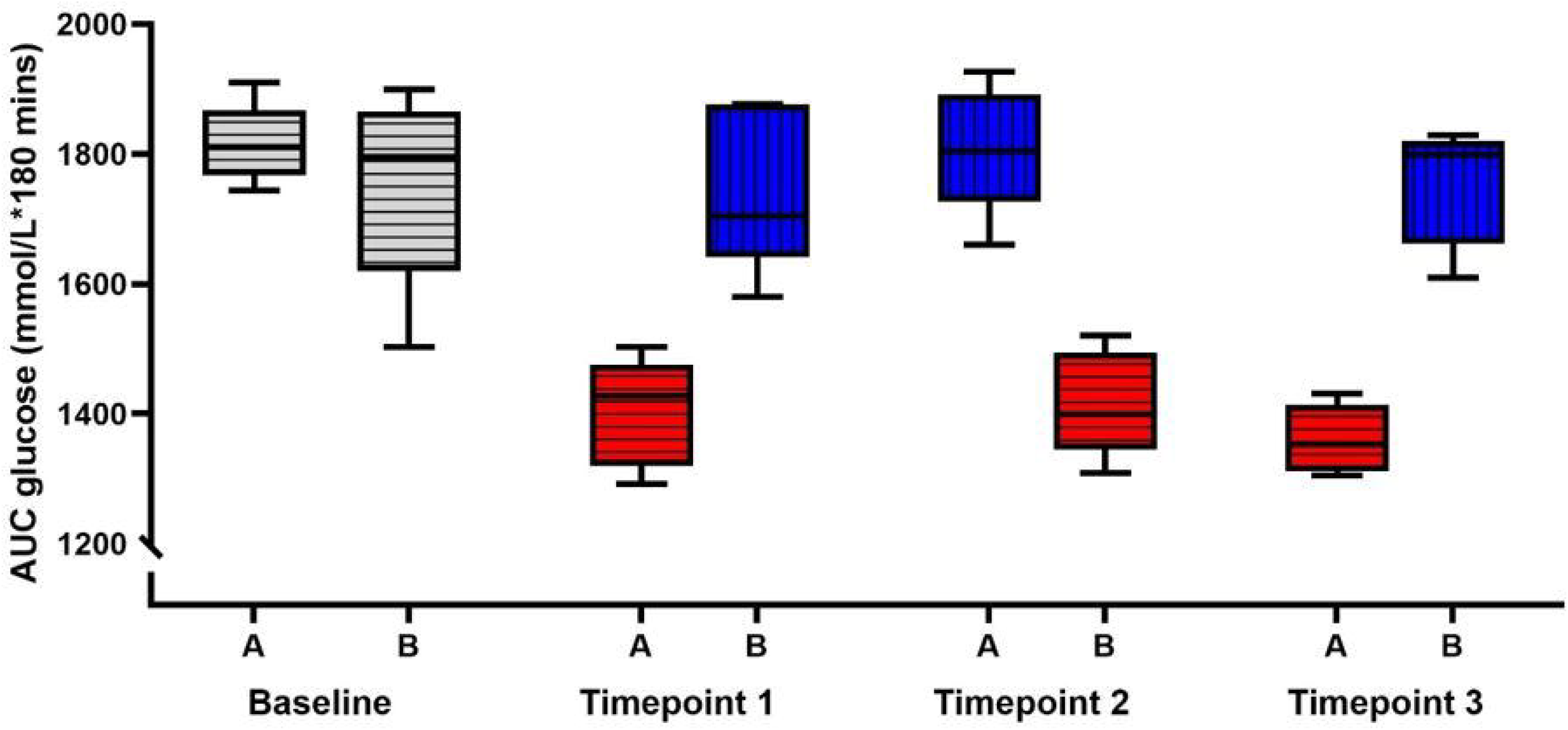

The daily checklist of foods (completion rate: 54% (20/37 days)) indicated people were having (mean ± SD) 6.7 ± 1.1 portions of carbohydrate foods and 3.1 ± 0.6 servings of protein foods on their usual diets; 3.4 ± 0.6 portions of carbohydrate foods and 6.1 ± 0.7 portions of protein foods during the high-protein condition and 3.5 ± 0.5 portions of carbohydrate foods and 2.9 ± 0.6 portions of protein foods during the low-protein condition.

The were no significant differences in weight at any timepoint between the high-protein and low-protein conditions (all p>0.75).

## Discussion

These findings add to the growing literature demonstrating a potent glucose-lowering effect of dietary protein in people with T2D, and extend our understanding findings by showing the effect remains without any change in carbohydrate. Mounting evidence suggests that while glucose-stimulated insulin secretion declines in T2D, amino-acid stimulated insulin secretion remains intact [7-9]. The insulinogenic effect of protein may therefore be of particular benefit for people with T2D seeking to lower their glucose without weight loss. These data also support emerging findings that modest reductions in carbohydrate - without reduction in calorie or increase in protein intake - do not lower blood glucose concentrations [4, 5].

Both fasting and postprandial glucose contribute to hyperglycemia in T2D [10]. In this study, the high-protein diet lowered both postprandial and fasting glucose, although the magnitude of effect was greater in the former and likely explained by the insulinogenic effect of elevated prandial circulating amino acids [8]. The hypoglycemic effect of high-protein, reduced carbohydrate diets on fasting glucose is less well understood. It’s possible that lowering of postprandial glucose reduces exposure of the beta cell to toxic concentrations of glucose (glucotoxicity) which indirectly leads to improvement in fasting glucose homeostasis [11].

However, amino acids also modulate hepatic lipid metabolism [3, 12-14] but whether a reduction in liver fat leads to lower endogenous glucose production is yet to be established. Future research should prioritize understanding how interactions between dietary protein and carbohydrate influence these pathways.

The cohort in this study were recently diagnosed with T2D and had glucose close to target at baseline. However, a high-protein, reduced-carbohydrate diet of comparable composition has previously been shown to lower glucose in people with much higher baseline glycaemia (HbA1c: 11.1%)[2]. Furthermore, since evidence from acute studies demonstrates beta-cell responsiveness to amino acids in people with T2D of long duration [7], future research should explore whether a high-protein, reduced-carbohydrate diet could be offered as a therapy for this population of patients.

The strengths of this study include the crossover design in which participants were exposed to one of the dietary conditions twice, and careful maintenance of bodyweight. While body weight did reduce this was in the order of expected weight loss with low carbohydrate diets [5], and did not differ between the high-protein and low-protein conditions. We did not use validated methods such as 24-hour recall or a food diary during this study, as these have been shown to be burdensome for participants. However, the checklist of foods suggested good compliance with the dietary conditions, and the changes in glycemia on the high protein condition reflect the magnitude of changes in glycaemia other studies in which this dietary pattern or similar dietary patterns have been used [3, 15].

In conclusion, this trial demonstrates that a high-protein, reduced-carbohydrate but not a low-protein, reduced carbohydrate diet reduces glycaemia in people with newly-diagnosed T2D independent of weight loss. Such a diet could help improve glycemia as a standalone or an adjunct to weight loss to help achieve optimum glucose concentrations.

## Supporting information

CONSORT diagram

## Data Availability

All data produced in the present study are available upon reasonable request to the authors

## Author Contributions

NG, NO and JB. were involved in the conception, design and conduct of the study. NG and VA carried out data extraction and analysis, with guidance from RS. NG is the guarantor of this work and, as such, had full access to all the data in the study and takes responsibility for the integrity of the data and the accuracy of the data analysis.

## Data Access and Availability

NG is the guarantor of this work and, as such, had full access to all the data in the study and takes responsibility for the integrity of the data and the accuracy of the data analysis.

The datasets generated during and/or analyzed in the current study are available from the corresponding author upon reasonable request.

