## Supplementary material for "A randomised crossover trial of high- versus low-protein diets on glycaemia in people with normal glucose tolerance (NGT) and type 2 diabetes (T2D)": CONSORT diagram

**Figure 1: CONSORT 2025 Flow Diagram**

Analysis

Analysed for primary outcome (n=20 )

Excluded from analysis (give reasons) (n=0 )

Analysed for primary outcome (n=20 )

Excluded from analysis (give reasons) (n=0 )

Allocated to low-protein (n=10)

Received allocated intervention (n=10)

Did not receive allocated intervention (n=0)

Allocated to high-protein (n=10)

Received allocated intervention (n=10)

Did not receive allocated intervention (change in circumstances) (n=0)

Allocated to high-protein (n=11)

Received allocated intervention (n=10)

Did not receive allocated intervention (change in circumstances) (n=1)

Third allocation

Allocated to low-protein (n=10)

Received allocated intervention (n=10)

Did not receive allocated intervention (n=0)

Second allocation

Allocated to low-protein (n=12)

Received allocated intervention (n=11)

Did not receive allocated intervention (change in circumstances) (n=1)

Allocated to high-protein (n=12)

Received allocated intervention (n=10)

Did not receive allocated intervention (change in circumstances) (n=2)

Allocation

Randomised (n=24 )

Excluded (n=102 )

Not meeting inclusion criteria (n=93)

Declined to participate (n=9)

Other reasons (n= )

Assessed for eligibility (n= 126)

Enrolment
